# Genetically proxied PDE5 inhibition and risk of dementia: a drug target Mendelian randomisation study

**DOI:** 10.1101/2024.02.15.24302874

**Authors:** Stephen O. Brennan, Alexander C. Tinworth

## Abstract

**Background:** Phosphodiesterase-5 (PDE5) inhibitors have gained interest as a potential treatment for dementia. However, current evidence is limited to observational and pre-clinical studies. This drug-target Mendelian Randomisation (MR) study aims to elucidate the on-target effects of pharmacological PDE5 inhibition on dementia subtypes, cognitive traits, and neuro-imaging phenotypes.

**Methods:** Two independent (r^2^ <0.001) blood pressure lowering variants from around the PDE5A locus were used in two-sample MR to assess the effect of genetically proxied PDE5 inhibition on risk of dementia subtypes, cognitive performance, and neuroimaging traits (cortical thickness, surface area and volume of white matter hyperintensities) in large-scale genomic consortia. The instrument’s predictive validity was assessed against erectile dysfunction and pulmonary arterial hypertension (PAH) as positive controls.

**Results:** Following correction for multiple comparisons, genetically proxied PDE5 inhibition was associated with lower odds of erectile dysfunction (OR 0.85, 95% CI 0.83-0.87) and PAH (OR 0.58, 95% CI 0.55-0.61), and higher odds of Alzheimer’s disease (OR 1.07, 95% CI 1.04-1.10), Lewy body dementia (OR 1.20, 95% CI 1.17-1.23) and vascular dementia (OR 1.04, 95% CI 1.02-1.07). Furthermore, genetically proxied PDE5 inhibition was associated with reduced cortical thickness (SD change -0.003, 95% CI -0.004, -0.002) and cognitive performance (SD change -0.010, 95% CI -0.013, -0.007), but not cortical surface area nor volume of white matter hyperintensities.

**Conclusion:** In contrast to evidence from observational studies, our findings indicate that inhibition of PDE5 is associated with a higher risk of dementia and an unfavourable neurocognitive profile. This risk should be further investigated prior to clinical trials of pharmacological PDE5 inhibition for the treatment and prevention of dementia.

## Introduction

Phosphodiesterase type 5 (PDE5) inhibitors such as Sildenafil (sold under the brand names Viagra, Revatio, and others) have recently gained attention as a potential treatment for neurodegenerative diseases, particularly Alzheimer’s disease (AD). AD is the most prevalent form of dementia worldwide and currently has few available disease-modifying and preventive therapies (1). Originally developed for erectile dysfunction (ED) and repurposed for pulmonary arterial hypertension (PAH), PDE5 inhibitors improve nitric-oxide-dependent vasodilation by preventing the breakdown of cyclic adenosine monophosphate (cAMP) and cyclic guanosine monophosphate (cGMP) in vascular smooth cells (2).

Several in vitro and pre-clinical studies have suggested that PDE5 inhibitors may offer protective effects against AD, Vascular dementia (VaD), and Lewy body dementia (LBD) (3). Additionally, three recent cohort studies have found that Sildenafil usage was associated with reduced risk of AD (4-6). However, despite efforts to minimise bias, these studies remain susceptible to inherent methodological limitations of observational research, such as reverse causality and residual confounding, which could account for the observed protective effects (7).

In the absence of trial evidence, genetic studies can offer insights into the on-target mechanisms of pharmacological agents (8). Mendelian randomisation (MR) utilises naturally occurring, randomly assorted genetic variation as instrumental variables to explore the causal relationship between an exposure and an outcome (9). Drug-target MR employs genetic variants that are predictive of the structure and function of a drug-target protein, which effectively act as a proxy for the exposure itself (10). These genetic variants are unaffected by reverse causation and environmental confounders and, on fulfilling the key assumptions of MR, can provide evidence of the causal effects of PDE5 inhibition on AD, VaD, and LBD (11). As an example, drug-target MR has already been used in the context of PDE5 inhibitors on the wellbeing and fertility of men (12).

This study aims to address the long-term effects of genetically proxied PDE5 inhibition on the risk of dementia subtypes by using genetic variants within the PDE5A locus associated with systolic blood pressure levels to mimic the effects of pharmacological PDE5 inhibition.

## Methods

### Study design

Two-sample cis-MR was used to assess the relationship between genetically proxied PDE5 inhibition and major dementia subtypes in European populations. We created a genetic instrument that proxies PDE5 inhibition by selecting independent variants from around the *PDE5A* gene locus that are strongly associated with systolic blood pressure, a downstream phenotype affected by PDE5 inhibitors. The predictive validity of this instrument was assessed against ED and PAH, before its use in primary analyses. Co-localisation analyses in a previous PDE5 inhibitor cis-MR study suggest that there is a high likelihood of a shared causal variant between systolic blood pressure and PDE5 protein quantitative trait loci and *PDE5A* expression quantitative trait loci, supporting the use of systolic blood pressure as an appropriate phenotype for constructing a genetic instrument for PDE5 inhibition (12). To determine the robustness of our findings, an alternative instrument was developed through a different selection methodology, which was previously applied in a recent MR study examining the impact of PDE5 inhibitors on male fertility (12). All contributing studies acquired appropriate informed consent from participants and ethical approval.

### Study populations/Genomic Consortia Systolic blood pressure

Summary statistics for systolic blood pressure were sourced from Evangelou et al.’s 2018 genome-wide association meta-analysis, encompassing 757,601 individuals of European descent from the International Consortium for Blood Pressure (ICBP) and the UK Biobank (UKB) (13). UKB is a large-scale prospective biomedical database comprising extensive health and genetic information on 500,000 participants (14).

### Erectile dysfunction and pulmonary arterial hypertension

Summary statistics for ED were obtained from Finngen (data freeze 9), and Bovijn et al.’s 2018 genome-wide association study of 6,175 self-reported, or physician-diagnosed cases of ED in UKB, the Estonian Genome Center of the University of Tartu (EGCUT) cohorts and the hospital-recruited Partners HealthCare Biobank (PHB) cohort (15). The FinnGen consortium contains data from nine Finnish biobanks and prospective cohort studies, cumulatively totalling 1,154 cases of ED and 318,913 controls ascertained through electronic linkage with nationwide registries. PAH summary statistics were derived from Finngen (125 cases) and Rhodes et al.’s GWAS of 2,085 patients from four international case-control studies, cumulatively totalling 2210 PAH cases and 174,581 controls (16).

### Dementia subtypes and cognitive phenotypes

Summary statistics for AD were obtained from Wightman et al.’s 2021 GWAS of clinically or autopsy-confirmed late-onset AD (17). To mitigate the effect of proxy cases and sample overlap, UKB data was excluded. Additionally, participant data from 23andMe was omitted due to privacy restrictions, yielding a total of 39,918 cases and 358,140 controls for inclusion in our analysis. Summary statistics for VaD (2,335 cases and 309,154 controls) were obtained from FinnGen (data freeze 9), and for LBD, from Chia et al.’s GWAS of pathologically or clinically diagnosed cases (2,591 cases and 6,618 controls) (18, 19).

### Cognitive performance, neuroimaging

Summary statistics for cognitive performance were derived from Lee et al., meta-analysis of UKB fluid intelligence and verbal-numerical reasoning scores, in addition to the Cognitive Genomics Consortium GWAS on neuropsychological test parameters (20). Associations between genetic variants and mean cortical thickness and surface area were sourced from a neuroimaging GWAS encompassing 33,709 individuals from 60 cohorts (21). Furthermore, data on the total volume of white matter hyperintensities ascertained from T1 and T2 flair images, were obtained from Smith et al.’s GWAS involving 32,114 participants from UKB (22).

### Instrumental variable selection

To satisfy the relevance and exclusion restriction criteria essential for valid MR analyses, we selected cis-variants associated with systolic blood pressure at genome-wide significance from within a 150kb flanking region of the *PDE5A* gene (hg19: chromosome 4:119494397-119628804). Variants were then clumped with an LD r^2^ threshold of <0.001 and a distance threshold of 250kb to select the variants with the strongest association with systolic blood pressure whilst minimising bias due to LD. This method selected two *cis*-variants associated with systolic blood pressure as described in Supplementary Table 1.

**Table 1.** Genetically proxied PDE5 inhibition and effects on cognitive performance and neuro-imaging phenotypes.

| Outcome | Mean (SD) | SD difference (95% CI) | P-value |
| --- | --- | --- | --- |
| Cognitive performance | Score 0 ± 1 | -0.010 (-0.013, -0.007) | <0.001 |
| Average cortical thickness | 2.45 ± 0.11mm | -0.003 (-0.004, -0.002) | <0.001 |
| Average cortical surface area | 169,647.43 ± 16,501.45mm<sup>2</sup> | 320.172 (25.464, 614.881) | 0.033 |
| White matter hyperintensities | -0.027 | -0.027 (-0.050, -0.005) | 0.016 |

Bonferroni corrected significance threshold: $p < 0.0036$ .

### Statistical analysis

In primary analyses, genetically proxied PDE5 inhibition was assessed against (1) established positive controls, (2) major dementia subtypes, and (3) measures of cognitive function in conjunction with neuroimaging phenotypes. In MR analyses, gene-exposure and gene-outcome data were harmonized, and the random-effects inverse-variance weighted (IVW) method was employed to evaluate the association between our genetic instrument and each outcome. To enhance statistical power, we conducted a fixed effect meta-analysis of the associations between genetically proxied phosphodiesterase 5 (PDE5) inhibition and each positive control from smaller non-overlapping data sources.

Results from all MR analyses are reported as odds ratios (OR) for binary outcomes or standard deviation (SD) difference for continuous outcomes, with 95% confidence intervals (CI) per SD lower in genetically predicted systolic blood pressure. When a genetic variant was absent in outcome summary statistics, a proxy variant with an LD r^2^ >0.95 was used in its place. The significance threshold was set at p=0.0036 after applying a Bonferroni correction for 14 statistical tests, involving two instruments and seven outcomes. Analyses were performed in R (v4.3,0; R Foundation for Statistical Computing) with the TwoSampleMR, MendelianRandomisation and metafor R packages.

### Sensitivity analyses

To further validate the robustness of our findings, we applied an alternate instrument selection method used in a recent PDE5 inhibitor drug-target MR study (23). Missense variants from within the *PD5EA* gene region associated with PDE5 protein levels and variants associated with *PDE5A* gene expression were ranked based on their p-value association with SBP and clumped with an LD threshold of r^2^ < 0.35 and a distance threshold of 10,000 kilobases (kB) which selected an instrumental variable containing four variants, as described in Supplementary Table 1.

MR sensitivity analyses including weighted median and weighted mode were used to assess for consistency of results, while Cochrane’s Q statistic and MR PRESSO were used to assess for heterogeneity and outliers, which can lead to horizontal pleiotropy and violation of the exclusion restriction assumption. Furthermore, we analysed our instrumental variants in PhenoScanner, a publicly available database of associations of human genotypes and phenotypes from large GWAS, to identify any associations these variants may have with other pleiotropic traits at p < 1×10^−5^ (24). This was followed with two-step cis-MR to adjust for any identified trait that could represent potential pleiotropy, which uses a two-step mediation approach to account for potential LD confounding or pleiotropic pathways (25).

## Results

### PDE5 inhibition and positive controls

Genetically proxied PDE5 inhibition was associated with 15% lower odds of ED (OR 0.85, 95% CI 0.83-0.87, p = <0.001) and 42% lower odds of PAH (OR 0.58, 95% CI 0.55-0.61, p = <0.001, Figure 1) with no evidence of heterogeneity (Supplementary Table 2). Our second instrument for genetically proxied PDE5 inhibition was strongly associated with lower odds of PAH, and directionally similar for ED (Supplementary Figure 1).

**Figure 1.**
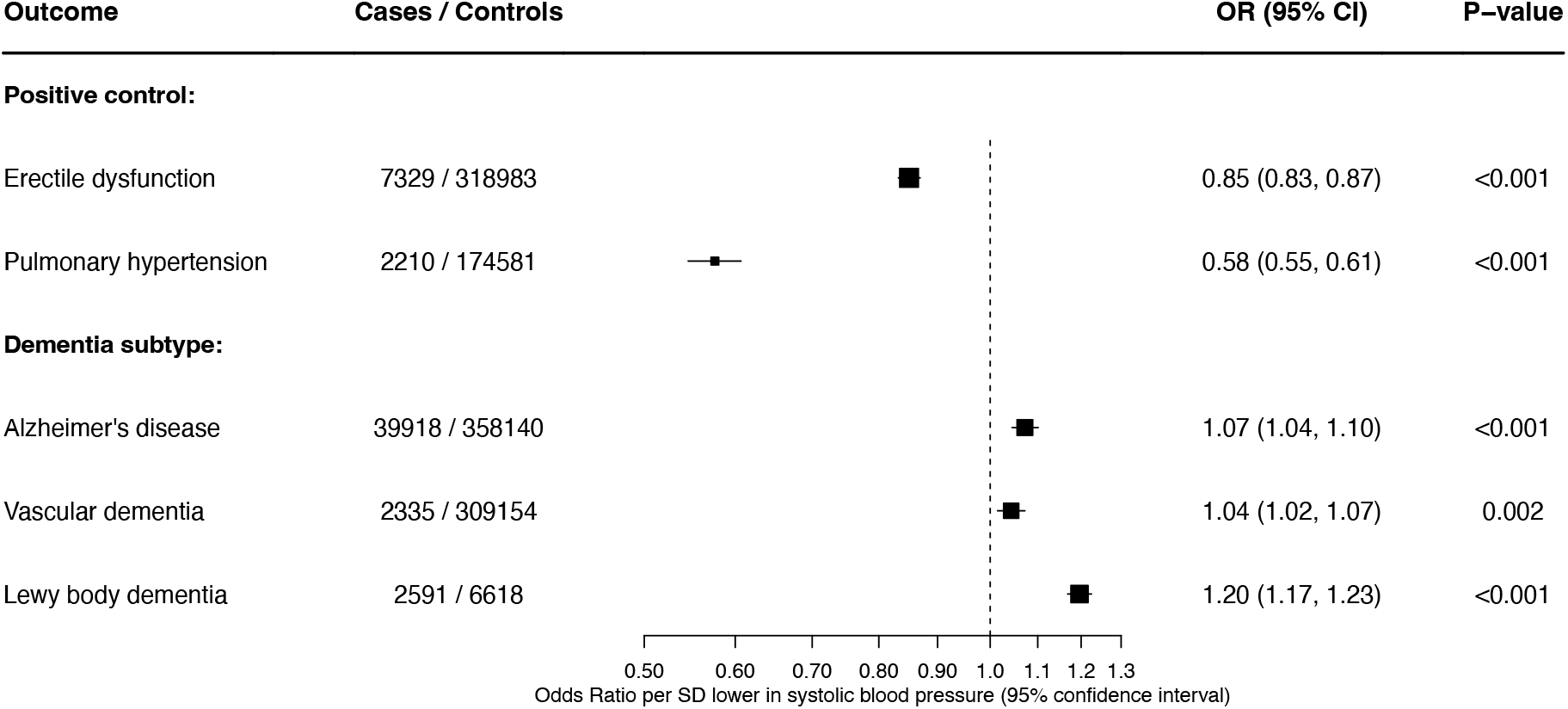
Genetically proxied PDE5 inhibition and associations with positive controls and dementia subtypes. Bonferroni corrected significance threshold: p < 0.0036.

### PDE5 inhibition and dementia subtypes

Genetically proxied PDE5 inhibition was associated with higher odds of AD (OR 1.07, 95% CI 1.04- 1.10, p = <0.001), and VaD (OR 1.04, 95% CI 1.02-1.07, p=0.002). Moreover, there was significantly higher odds of LBD, with each SD lower in genetically predicted systolic pressure associated with 20% higher odds of disease (OR 1.20, 95% CI 1.17-1.23, p = <0.001, Figure 1).

Results were directionally similar when using our second instrument for genetically proxied PDE5 inhibition (Supplementary Figure 1), and when using weighted median, weighted mode and MR-PRESSO methods with no evidence of heterogeneity (Supplementary Table 2). However, the second instrument was not significantly associated with LBD (OR 1.35, 95% CI 0.97-1.90, p = 0.08) (Supplementary Figure 1).

OR: odds ratio, CI: confidence interval, SD: standard deviation. Bonferroni corrected significance threshold: p < 0.0036.

### PDE5 inhibition and cognitive performance and structural neuro-imaging outcomes

Genetically proxied PDE5 inhibition was associated with lower cognitive performance (SD change - 0.010, 95% CI -0.013, -0.007, p = <0.001) and lower mean cortical thickness (SD change -0.003, 95% CI 0.004, -0.002, p = 0.002, Table 1). There was no association between genetically proxied PDE5 inhibition and average cortical surface area or white matter hyperintensities. Results were directionally similar when using weighted median, weighted mode and MR-PRESSO methods with no evidence of heterogeneity (Supplementary Table 2). Our second instrument for genetically proxied PDE5 inhibition was associated with decreased cognitive performance and reduced volume of white matter hyperintensities. However, these associations did not remain significant after adjusting for multiple comparisons (Supplementary Table 2).

### Pleiotropic effects

Through lookups on PhenoScanner, seven traits were identified that were associated (p < 1×10^−5^) with at least one of the variants included in the genetic instrument (Supplementary Table 3). In two-step cis-MR, none of the identified traits significantly altered the association between genetically proxied PDE5 inhibition and AD or VaD (Supplementary Table 4).

## Discussion

This study utilised genetic epidemiology to provide novel insights into the relationship between PDE5 inhibition and dementia. Leveraging data from large-scale genomic consortia and following adjustment for multiple testing, genetically proxied PDE5 inhibition was associated with increased risk of AD, LBD and VaD when using our primary instrument, suggesting a potential causal relationship. In a further triangulation of findings, genetically proxied PDE5 inhibition was associated with reduced cognitive performance and cortical thickness. When using the secondary instrument, there was a consistent increase in risk of AD and VaD, but not LBD, and when adjusting for multiple testing, the secondary instrument did not associate with neuroimaging phenotypes or cognitive performance. However, the lack of a significant association with ED in the secondary instrument suggests a weakness as a valid proxy of PDE5 inhibition compared to the primary instrument, which was constructed as a conventional instrumental variable for investigating drug-target MR (10).

Several recent observational studies have assessed the impact of PDE5 inhibitors on risk of AD (4-6). Adesuyan et al. found that PDE5 inhibitor prescription was associated with an 18% lower risk of disease in men with ED, while Fang et al., found 69% lower risk of AD with prescription of a PDE5i (4, 6). In contrast, Desai et al. found no association between PDE5 inhibitor prescription and AD in people with PAH (26). Our analyses, the first genetic epidemiological study thus far of PDE5 inhibition and dementia, contrast with these previous findings. Dementia’s extended prodromal phase can lead to reverse causality, where the development of dementia affects an exposure, as seen with the protective effect observed with a higher late-life body mass index, which disappears when early years of follow-up are excluded (27, 28). Similarly, PDE5 inhibitors are more likely to be prescribed in healthier individuals not yet affected by the prodromal period of dementia. This selective prescription can introduce bias in studies that fail to adequately exclude the early years of follow-up. This phenomenon was observed in Adesuyan et al.’s study, where results lost significance following the exclusion of the first three years of follow-up (6).

Furthermore, residual confounding is a common bias seen in observational analyses, and while certain measured confounders can be adjusted for statistically, it is challenging to account for unmeasured confounders (29). All three studies fail to adjust for education and *apolipoprotein E* status, which are well-described risk factors for the development of dementia that may also influence the prescription of phosphodiesterase inhibitors (30, 31).

Mechanistic support for the benefit of PDE5 inhibition is currently limited to preclinical animal studies (32-34). One of the major theories hypothesized to explain the potential benefits of PDE5 inhibition on the development of dementia is through vasodilation and improvements in cerebral blood flow which leads to improved perfusion of downstream brain tissue (35, 36). However, the PASTIS randomised controlled trial (RCT) did not show a difference in cerebral blood flow with taldafil compared to placebo in older people with symptomatic small vessel disease, a patient population at high risk for the development of VaD (37). Similarly, another RCT found that PDE5 inhibition had no positive effect on behavioural outcomes, along with negative effects on cerebral haemodynamics (38). Sustained PDE5 inhibition may counterintuitively reduce cAMP levels due to activation of PDE2 from chronically increased cGMP concentration, resulting in the harmful effects seen in those with genetic variants mimicking PDE5 inhibition (39). This may have been brought to light by the MR study design, which estimates a lifetime risk due to inherited differences in PDE5 inhibition compared to shorter-term use following diagnosis of ED or PAH. How relevant this is to pharmacological PDE5 inhibition compared its genetic proxy requires further investigation, but the aforementioned RCT showing negative effects on verbal fluency, memory tests, and prefrontal cortex activation suggests validity to the concerns highlighted in this study (38).

### Strengths

The primary strength of this study lies in the development and implementation of a genetic instrument that reliably proxies PDE5 inhibition. Unlike observational studies, this approach is robust to confounding factors and reverse causation. The validity of our results is further supported by successfully replicating the known pharmacological effects of PDE5 inhibitors on ED and PAH.

Furthermore, case ascertainment of each dementia subtype was based on clinically diagnosed or pathologically confirmed cases, mitigating the potential bias of including proxy dementia cases based on family history with no guarantee of clinical manifestation. The consistency of results across two genetic instruments and MR methods, which adjust for potential pleiotropy, along with the triangulation of evidence from neuroimaging data and cognitive tests, strengthens confidence in our primary findings. Finally, on top of addressing potential pleiotropy statistically, the findings from PhenoScanner combined with subsequent two-step cis-MR further reduce the likelihood of pleiotropy influencing the study results.

### Limitations

Several limitations should be considered when interpreting these results. Data was not available on the proportion of cases clinically diagnosed through imaging and cerebrospinal fluid biomarkers in large genetic consortia which raises potential concerns about case misclassification. Effect sizes derived from drug-target MR studies reflect lifelong perturbation of the drug target rather than the short-term treatment duration typically seen in clinical practice, potentially leading to significant differences in the magnitude of benefit (10). Similarly, there may be temporal interactions between PDE5 inhibitors and neurocognitive disorders that may not become clinically relevant until the condition has reached a certain level of pathophysiology (40). Summary statistics for dementia are predominantly available in those of European ancestry, limiting our ability to assess the effect of genetically proxied PDE5 inhibition across ancestries.

## Conclusion

This drug-target MR study has explored the effect of genetically proxied PDE5 inhibition on the risk of dementia and neurocognitive phenotypes. Findings indicate that PDE5 inhibition may be causally associated with a higher risk of AD, VaD and LBD. Further studies exploring potential mediating pathways of harmful effects are warranted prior to clinical trials of pharmacological PDE5 inhibition in the treatment and prevention of dementia.

## Data availability

This study utilises publicly available, summary-level GWAS data that can be accessed through the GWAS Catalogue (https://www.ebi.ac.uk/gwas/home) or specific cohort portals, including FinnGen (https://www.finngen.fi/en/access_results), the Social Science Genetic Association Consortium (https://www.thessgac.org/), and ENIGMA (https://enigma.ini.usc.edu/).

## Supporting information

Supplementary Methods and Results

