## Supplementary Methods and Results for "Genetically proxied PDE5 inhibition and risk of dementia: a drug target Mendelian randomisation study"

**Supplemental online content**

### Supplementary Methods

*Study design*

Mendelian randomisation (MR) uses genetic variants associated with an exposure of interest, commonly obtained from genome-wide association studies (GWAS), to act as an instrumental variable when exploring causal relationships between that exposure and an outcome. Due to Mendel's laws of segregation and independent assortment, genetic variants are randomly distributed throughout a population, and therefore mimic the randomisation seen in randomised controlled trials (RCT) (1). Additionally, due to being assigned at conception, these instrumental variables do not suffer from reverse causation, a common bias seen in observational studies.

Estimates from MR analyses can be considered causal if three assumptions are met: (i) relevance, where the genetic instrument is associated with the exposure of interest, (ii) exchangeability, where the instrument is not associated with confounders of the exposure-outcome relationship, and (iii) the exclusion restriction assumption, where the instrument does not affect the outcome through a pathway separate to that of the exposure. The relevance assumption is often satisfied through generation of F-statistics, where an F statistic above 10 is considered sufficient to avoid weak instrument bias, but the other two assumptions can only be partially addressed (2). MR is continually updated with new methodologies that can mitigate potential violations of the three assumptions, with careful consideration required for their applicability depending on the research question.

Recently, the scope of MR (Mendelian Randomization) has broadened to include the investigation of drug targets. Most existing drugs target specific proteins and the assessment of genetic variants from around the gene loci of these proteins can be used to assess the effect of perturbing the drug target protein on outcomes of interest. Genetic variants within or near the locus of the gene encoding the drug target protein of interest are known as *cis*-variants, and these can be used in MR to investigate current or novel drug targets (3). Drug target MR is still subject to the same assumptions as conventional MR, with additional focus on validation steps that ensure the genetic instrument captures the effect of its pharmacological intervention, often by checking for associations with positive control outcomes (4). Another important consideration is that the investigated perturbation is measured over an individual's lifetime, compared to an RCT where the intervention is over a shorter period (5).

*Data sources*

*Systolic blood pressure*

Mean systolic blood pressure was derived from two measurements in UK Biobank (UKB) (automated or manual) when available or for a small subset through a single measurement (N=413). Following quality control, 458,577 individuals from UKB were included, and blood pressure medications were adjusted for where appropriate. For the GWAS, association analyses were performed using a linear mixed model that adjusted for age, age$^2$, body mass index, and the optional inclusion of covariates that controlled for population stratification. For the International Consortium for Blood Pressure (ICBP) GWAS, post-quality control data from 54 studies comprising 150,134 individuals was combined with 23 independent studies of 148,890 individuals in total resulting in a dataset of 299,024 individuals of European ancestry. Summary data from the UKB and ICBP were combined using a fixed-effects inverse variance weighted meta-analysis (6).

*Pulmonary arterial hypertension*

Pulmonary arterial hypertension was defined across the four included studies through haemodynamic criteria according to international guidelines, namely, an increase in mean pulmonary arterial pressure by ≥25 at rest through right heart catheter assessment (7). Individuals were included if they were unrelated and cases resulting from autoimmune disease were excluded. SNP associations were assessed through logistic regression adjusting for sex, read length chemistry, and principle components (number depended on each individual study). Findings were cross-validated across all four studies using the inverse variance-weighted fixed effects meta-analysis approach (8).

*Dementia subtypes*

Alzheimer's disease (AD) cases were obtained from Wightman et al.'s GWAS of late-onset AD from thirteen cohort studies. However, for the purposes of this study, two datasets (UKB and 23andMe) were excluded due to inclusion of proxy cases, sample overlap, and privacy restrictions. Case definition varied slightly between the datasets but were either registered to ICD10 codes F00 and G30, defined from medical assessment, or neurological evaluation, and controls were considered healthy with no prior diagnosis of AD (9).

For vascular dementia, cases in FinnGen (a cohort study with over 300,000 genotyped individuals of Finnish ancestry living in Finland) were recorded as ICD-10 F01, and Lewy body dementia was defined by Chia et al. as clinically probable (802 cases) or through autopsy confirmation (1,789 cases) (10). Controls were selected based on a lack of evidence of cognitive decline or neurological deficits on neurological examination (11).

*Cognitive performance and neuroimaging*

Summary statistics for cognitive performance were obtained from Lee et. al.'s genome-wide association meta-analysis of general cognitive ability in 32,298 European-ancestry individuals from the COGENT consortium and cognitive performance in 222,532 UKB participants (12). In COGENT, 35 participating cohorts assessed cognitive performance through three or more neuropsychological tests, while in UKB, cognitive performance was assessed through a test of verbal-numerical reasoning. Summary statistics for cortical surface area and average cortical thickness were obtained from Grasby et. al.'s genome-wide association meta-analysis of brain magnetic resonance imaging (MRI) data from 33,992 participants across 50 cohorts (13). Summary statistics for the volume of white matter hyperintensities were obtained from Smith et al.'s GWAS, which included 33,224 UKB participants who underwent multimodal brain imaging (14). This imaging-derived phenotype was assessed using T2-weighted FLAIR structural imaging.

*Two-step cis-Mendelian randomization*

All summary statistics used for the two-step *cis*-MR portion of the analyses were obtained from the Integrative Epidemiology Unit (IEU) OpenGWAS project in individuals of European ancestry. Summary statistics for plateletcrit (n = 164,339, IEU OpenGWAS ID: ebi-a-GCST004607), myeloid white cell count (n = 169,219, IEU OpenGWAS ID: ebi-a-GCST004626), granulocyte count (n = 169,822, IEU OpenGWAS ID: 169,822), and sum basophil and neutrophil count (n = 170,143, IEU OpenGWAS ID: ebi-a-GCST004620) were obtained from a GWAS of the UKB and INTERVAL studies (15). Coronary artery disease summary statistics (cases = 122,733, controls = 424,528, IEU

OpenGWAS ID: ebi-a-GCST005195) was obtained from a GWAS of UKB and CARDIoGRAMplusC4D (16). White blood cell count summary statistics (n = 563,946, IEU OpenGWAS ID: ieu-b-30) were obtained from many consortia that were manually curated by the IEU. Impedance of right leg (n = 454,863, IEU OpenGWAS ID: ukb-b-7376), impedance of left leg (n = 454,857, IEU OpenGWAS ID: ukb-b-14068), impedance of right arm (n = 454,826, IEU OpenGWAS ID: ukb-b-7859), impedance of left arm (n = 454,850, IEU OpenGWAS ID: ukb-b-19379), impedance of whole body (n = 454,840, IEU OpenGWAS ID: ukb-b-19921), standing height (n = 461,950, IEU OpenGWAS ID: ukb-b-10787), body mass index (n = 461,460, IEU OpenGWAS ID: ukb-b-19953), and platelet count (n = 350,474, IEU OpenGWAS ID: ukd-d-30080_irnt) were extracted from various GWAS conducted on UKB (17, 18).

*Sensitivity analysis*

A second genetic instrument used in a previous study investigating genetically proxied PDE5 inhibition, was developed to assess the consistency of results across different instrumental variable selection methods (19). This involved selecting GWAS significant ($p<5x10^{-8}$) missense pQTLs variants or eQTLs in the *PDE5A* gene locus that were ranked in order of P-values and selected based on a linkage disequilibrium threshold of $r^2<0.35$ and distance threshold of 10,000 kilobases. This selection methodology increases statistical power through the selection of mildly correlated rather than strictly independent variants that influence both the expression and levels of PDE5, with the thresholds selected to balance the statistical bias induced by including too highly correlated variants. In our study, this method doubled the number of included SNPs from two to four (Supplementary table 1).

Four additional MR methods were adopted to assess the consistency of results. The weighted median and mode methods assume some genetic variants are valid but are more robust to potential pleiotropic outliers than the inverse variance-weighted and Egger methods (20). Additionally, the MR-PRESSO method was also adopted which removes genetic variants from the analysis that have substantial differences in their estimate compared to other variants and performs the inverse variance-weighted method on the remaining variants (4). Heterogeneity between variants was assessed through Cochran's Q statistic which follows an $X^2$ distribution with degrees of freedom defined as n number of SNPs minus 1. A P-value <0.05 indicates a violation of the MR assumptions across some or all of the genetic variants (21). Finally, traits identified by SNP lookups on PhenoScanner were adjusted for in two-step cis-MR, a method developed to address potential pleiotropy and linkage disequilibrium confounding. The method relies on splitting the MR estimate into an estimate contributed from the variant through the exposure, and an estimate contributed from the variant through the confounder, the latter is subtracted from the former to leave a theoretically unbiased overall estimate (19).

### Supplementary results

**Supplementary Figure 1.** Alternative genetic proxy for PDE5 inhibition and associations with positive controls and dementia subtypes

| Outcome | Cases / Controls | OR (95% CI) | P−value |
|---|---|---|---|
| **Positive controls:** | | | |
| Erectile dysfunction | 7329 / 318983 | 0.91 (0.82, 1.01) | 0.076 |
| Pulmonary hypertension | 2210 / 174581 | 0.80 (0.68, 0.94) | 0.007 |
| **Dementia subtypes**: | | | |
| Alzheimer's disease | 39918 / 358140 | 1.08 (1.03, 1.13) | <0.001 |
| Vascular dementia | 2335 / 309154 | 1.20 (1.05, 1.37) | 0.003 |
| Lewy body dementia | 2591 / 6618 | 1.29 (0.98, 1.71) | 0.07 |

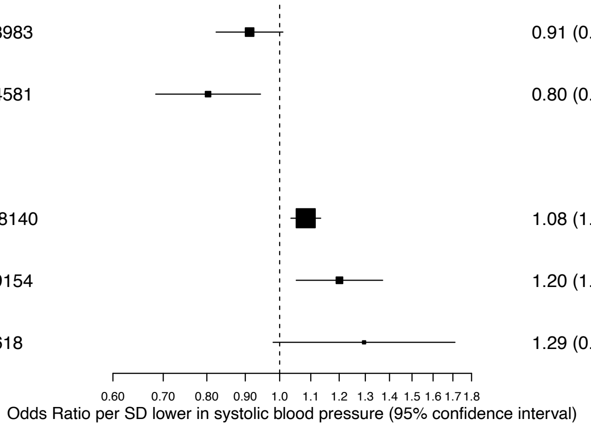

OR: odds ratio, CI: confidence interval, SD: standard deviation. Bonferroni corrected significance threshold: p < 0.0036.

**STROBE-MR Checklist**

| Item No. | Section | | Checklist item | Page No. | Relevant text from manuscript |
|---|---|---|---|---|---|
| 1 | **TITLE and ABSTRACT** | | Indicate Mendelian randomization (MR) as the study's design in the title and/or the abstract if that is a main purpose of the study | 1 | Inserted title and abstract |
| | **INTRODUCTION** | | | | |
| 2 | **Background** | | Explain the scientific background and rationale for the reported study. What is the exposure? Is a potential causal relationship between exposure and outcome plausible? Justify why MR is a helpful method to address the study question | 2 | Introduction |
| 3 | **Objectives** | | State specific objectives clearly, including pre-specified causal hypotheses (if any). State that MR is a method that, under specific assumptions, intends to estimate causal effects | 2 | Introduction – final paragraph |
| | **METHODS** | | | | |
| 4 | **Study design and data sources** | | Present key elements of the study design early in the article. Consider including a table listing sources of data for all phases of the study. For each data source contributing to the analysis, describe the following: | | |
| | | a) | Setting: Describe the study design and the underlying population, if possible. Describe the setting, locations, and relevant dates, including periods of recruitment, exposure, follow-up, and data collection, when available. | 3 | Study design |
| | | b) | Participants: Give the eligibility criteria, and the sources and methods of selection of participants. Report the sample size, and whether any power or sample size calculations were carried out prior to the main analysis | 3-4 | Study populations/Genetic Consortia |
| | | c) | Describe measurement, quality control and selection of genetic variants | 4 | Instrument creation |
| | | d) | For each exposure, outcome, and other relevant variables, describe methods of assessment and diagnostic criteria for diseases | 3/4 | Study populations/Genetic Consortia. Instrument creation, Supplementary section: Data sources |
| | | e) | Provide details of ethics committee approval and participant informed consent, if relevant | 3 | Study design |

| | | | | | |
|---|---|---|---|---|---|
| 5 | **Assumptions** | | Explicitly state the three core IV assumptions for the main analysis (relevance, independence and exclusion restriction) as well assumptions for any additional or sensitivity analysis | Supplementary methods | |
| 6 | **Statistical methods: main analysis** | | Describe statistical methods and statistics used | | |
| | | a) | Describe how quantitative variables were handled in the analyses (i.e., scale, units, model) | 4 | Statistical analysis |
| | | b) | Describe how genetic variants were handled in the analyses and, if applicable, how their weights were selected | 4-5 | Statistical analysis |
| | | c) | Describe the MR estimator (e.g. two-stage least squares, Wald ratio) and related statistics. Detail the included covariates and, in case of two-sample MR, whether the same covariate set was used for adjustment in the two samples | 4 | Statistical analysis |
| | | d) | Explain how missing data were addressed | NA | NA |
| | | e) | If applicable, indicate how multiple testing was addressed | 4 | Statistical analysis |
| 7 | **Assessment of assumptions** | | Describe any methods or prior knowledge used to assess the assumptions or justify their validity | 5 | Sensitivity analysis |
| 8 | **Sensitivity analyses and additional analyses** | | Describe any sensitivity analyses or additional analyses performed (e.g. comparison of effect estimates from different approaches, independent replication, bias analytic techniques, validation of instruments, simulations) | 5 | Sensitivity analysis |
| 9 | **Software and pre-registration** | | | | |
| | | a) | Name statistical software and package(s), including version and settings used | 4 | Statistical analysis |
| | | b) | State whether the study protocol and details were pre-registered (as well as when and where) | NA | NA |
| | **RESULTS** | | | | |
| 10 | **Descriptive data** | | | | |

| | | | | | |
|---|---|---|---|---|---|
| | | a) | Report the numbers of individuals at each stage of included studies and reasons for exclusion. Consider use of a flow diagram | Figure 1 | |
| | | b) | Report summary statistics for phenotypic exposure(s), outcome(s), and other relevant variables (e.g. means, SDs, proportions) | NA | |
| | | c) | If the data sources include meta-analyses of previous studies, provide the assessments of heterogeneity across these studies | NA | |
| | | d) | For two-sample MR: | | |
| | | | i. Provide justification of the similarity of the genetic variant-exposure associations between the exposure and outcome samples | NA | |
| | | | ii. Provide information on the number of individuals who overlap between the exposure and outcome studies | 3 | Study populations/Genetic Consortia |
| 11 | **Main results** | | | | |
| | | a) | Report the associations between genetic variant and exposure, and between genetic variant and outcome, preferably on an interpretable scale | Supplementary Tables | Supplementary Table 1 |
| | | b) | Report MR estimates of the relationship between exposure and outcome, and the measures of uncertainty from the MR analysis, on an interpretable scale, such as odds ratio or relative risk per SD difference | 5-6 | Results |
| | | c) | If relevant, consider translating estimates of relative risk into absolute risk for a meaningful time period | N/A | |
| | | d) | Consider plots to visualize results (e.g. forest plot, scatterplot of associations between genetic variants and outcome versus between genetic variants and exposure) | Figure 1 | |
| 12 | **Assessment of assumptions** | | | | |
| | | a) | Report the assessment of the validity of the assumptions | 5-6 | Results |
| | | b) | Report any additional statistics (e.g., assessments of heterogeneity across genetic variants, such as $I^2$, Q statistic or E-value) | 5-6 | Results |
| 13 | **Sensitivity analyses and** | | | | |

| | | | | | |
|---|---|---|---|---|---|
| | **additional analyses** | | | | |
| | | a) | Report any sensitivity analyses to assess the robustness of the main results to violations of the assumptions | 5-7 | Results |
| | | b) | Report results from other sensitivity analyses or additional analyses | NA | |
| | | c) | Report any assessment of direction of causal relationship (e.g., bidirectional MR) | NA | |
| | | d) | When relevant, report and compare with estimates from non-MR analyses | NA | |
| | | e) | Consider additional plots to visualize results (e.g., leave-one-out analyses) | Supplementary Tables | Supplementary Table 2, 3, 4 |
| | **DISCUSSION** | | | | |
| 14 | **Key results** | | Summarize key results with reference to study objectives | 7 | Discussion |
| 15 | **Limitations** | | Discuss limitations of the study, taking into account the validity of the IV assumptions, other sources of potential bias, and imprecision. Discuss both direction and magnitude of any potential bias and any efforts to address them | 8 | Limitations |
| 16 | **Interpretation** | | | | |
| | | a) | Meaning: Give a cautious overall interpretation of results in the context of their limitations and in comparison with other studies | 7 | Discussion |
| | | b) | Mechanism: Discuss underlying biological mechanisms that could drive a potential causal relationship between the investigated exposure and the outcome, and whether the gene-environment equivalence assumption is reasonable. Use causal language carefully, clarifying that IV estimates may provide causal effects only under certain assumptions | 7 | Discussion |
| | | c) | Clinical relevance: Discuss whether the results have clinical or public policy relevance, and to what extent they inform effect sizes of possible interventions | 7 | Discussion |
| 17 | **Generalizability** | | Discuss the generalizability of the study results (a) to other populations, (b) across other exposure periods/timings, and (c) across other levels of exposure | 8 | Limitations |
| | **OTHER INFORMATION** | | | | |

| | | | |
|---|---|---|---|
| 18 | **Funding** | Describe sources of funding and the role of funders in the present study and, if applicable, sources of funding for the databases and original study or studies on which the present study is based | 9 |
| 19 | **Data and data sharing** | Provide the data used to perform all analyses or report where and how the data can be accessed, and reference these sources in the article. Provide the statistical code needed to reproduce the results in the article, or report whether the code is publicly accessible and if so, where | 9 |
| 20 | **Conflicts of Interest** | All authors should declare all potential conflicts of interest | 9 |
